# Accelerometer-derived sleep onset timing and cardiovascular disease incidence: a UK Biobank cohort study

**DOI:** 10.1101/2021.06.23.21259390

**Authors:** Shahram Nikbakhtian, Angus B Reed, Bernard Dillon Obika, Davide Morelli, Adam C Cunningham, Mert Aral, David Plans

## Abstract

**Aims:** Growing evidence suggests that sleep quality is associated with cardiovascular risk. However, research in this area often relies upon recollection dependant questionnaires or diaries. Accelerometers provide an alternative tool for deriving sleep parameters measuring sleep patterns objectively. This study examines the associations between accelerometer derived sleep onset timing and cardiovascular disease (CVD).

**Methods and Results:** We derived sleep onset and waking up time from accelerometer data collected from 103,712 UK Biobank participants over a period of seven days. From this, we examined the association between sleep onset timing and CVD incidence using a series of Cox proportional hazards models. 3172 cases of CVD were reported during a mean follow-up period of 5·7 (±0·49) years. An age- and sex-controlled base analysis found that sleep onset time of 10:00pm-10:59pm was associated with the lowest CVD incidence. A fully adjusted model, additionally controlling for sleep duration, sleep irregularity, and established CVD risk factors, was unable to eliminate this association, producing hazard ratios of 1·24 (95% CI, 1·10–1·39; *p*<0·005), 1·12 (1·01–1·25; *p=*0·04), and 1·25 (1·02–1·52; *p=*0·03) for sleep onset <10:00pm, 11:00pm-11:59pm, and ≥12:00am, respectively, compared to 10:00pm-10:59pm. Importantly, sensitivity analyses revealed this association was stronger in females, with only sleep onset <10:00pm significant for males.

**Conclusions:** Our findings suggest an independent relationship between sleep onset timing and risk of developing CVD, particularly for women. We also demonstrate the potential utility of collecting information about sleep parameters via accelerometry-capable wearable devices, which may serve as novel cardiovascular risk indicators.

**Translational Perspective:** Abnormal sleep timing is a feature of poor-quality sleep and is likely to be associated with cardiovascular diseases. Accelerometer-derived measures of sleep quality, particularly sleep timing, may be associated with greater vulnerability to cardiovascular disease, particularly in women. Onset of sleep timing may constitute a novel cardiovascular risk factor and target for preventative intervention. Wearable devices equipped with accelerometers may be useful in assessing sleep related cardiovascular disease risk factors as they would allow passive measurement of sleep parameters, such as sleep timing.

## Introduction

Cardiovascular disease (CVD) continues to be the most significant cause of mortality worldwide, with an estimated 18·6 million deaths each year.^1^ Primary prevention of CVD has become a mainstay of addressing this considerable global challenge. The majority of clinical efforts are directed towards minimising modifiable risk factors through lifestyle changes and medical intervention. Traditional modifiable risk factors are well defined, including hypertension, obesity, diabetes, smoking, and hypercholesterolaemia.^2^ These have all been shown to improve with lifestyle changes. However, poor population-level adherence to lifestyle advice is a considerable challenge to preventive efforts.^3^

Patients require the personal capability, opportunity, and motivation to adopt and sustain lifestyle changes.^4^ Aside from individual hesitancy towards behavioural change, factors such as socio-economic constraints can impact capability and opportunity regarding exercise, dietary changes, and smoking cessation, limiting the impact of this public health guidance. Thus, identifying additional lifestyle risk factors that require effortless adherence to modify is of significant value to public health.

Emerging evidence suggests that disturbance of circadian rhythm, which incorporates essential physiological and humoral functions in a 24-hour cycle, could be an understudied risk factor for CVD. Prolonged misalignment of circadian rhythms has been associated with elevated blood pressures, reduced sleep quality, and increased risk for cardiovascular disorders.^5–7^ It is known that CVD events, such as stroke, cardiac arrest, and myocardial infarction, are more common during the early morning when heart rate, blood pressure, and cortisol and other hormone levels rise.^8,9^ Disruption of circadian rhythm may also stimulate atherosclerosis, providing a possible biological mechanism for increased cardiovascular risk.^10,11^ Additionally, such disruption may promote CVD in a sexually dimorphic manner.^12^

While impaired sleep quality is a significant contributor to circadian rhythm disturbance, the evidence regarding sleep interventions and their effect on cardiovascular events is lacking.^13^ Most previous studies investigating sleep disruption have had limitations, such as relying on subjective reporting of sleep, small or selective samples, and minimal adjustment for confounding factors. Sleep quality can generally be measured using three parameters: sleep duration, sleep irregularity, and sleep timing. Sleep duration is the total time spent asleep during the night; sleep timing corresponds to the timing of going to sleep and subsequent waking; while sleep irregularity has been defined as the variation in both sleep duration and timing displayed by an individual over a period of time.^14^

Among these three parameters, the relationship between CVD and sleep duration has been widely studied, while irregularity and timing have received much less attention. The influence of sleep irregularity on CVD risk has been investigated in recent research,^14^ while only one study has examined the relationship between sleep timing and congestive heart failure using sleep habit questionnaires.^15^

An analysis of Danish nurses found that proportionally fewer day shifts resulted in increased risk of cardiovascular-related mortality, with night shifts conferring greater risk than evening shifts.^16^ Understanding which factors are contributing to such observations is of high value to managing population-level health. Sleep timing represents a potential explanation for shift work-associated cardiovascular risk and, thereby, an avenue towards a novel cardiovascular risk factor, which would require minimal intervention to mitigate with lifestyle adjustment. Currently, there is a lack of research exploiting objectively measured sleep parameters in large population-based samples; hence our investigation aims to assess the association between sleep timing and CVD risk through accelerometry data in the large, comprehensively characterised UK Biobank (UKB) cohort.

## Methods

The UKB cohort consists of more than 500,000 participants aged 37-73, recruited between 2006 and 2010. All participants completed a range of demographic, lifestyle, health, and physical assessments and questionnaires. A subsample (103,712) also provided accelerometer data using a wrist-worn accelerometer.

Participants provided informed consent, and the UKB study received ethical approval from the North West Multi-Centre Research Ethics Committee (REC reference: 16/NW/0382) and was conducted in accordance with the principles of the Declaration of Helsinki. The current analyses were conducted under UKB application number 55668.

### Accelerometry data collection and pre-processing

Between 2013 and 2015, 103,712 participants enrolled in the UKB accepted the invitation to wear an accelerometer for seven days and provided accelerometer data. The participants who accepted wore an Axivity AX3 triaxial accelerometer (https://axivity.com/product/ax3) on their dominant wrist and continued with normal daily activities. Over seven days, participants’ activity was recorded by the accelerometer. The accelerometer data was pre-processed by the UKB accelerometer expert working group.^17^ Details of data collection and pre-processing can be found at http://biobank.ctsu.ox.ac.uk/crystal/refer.cgi?id=131600.

### Accelerometer data processing

We derived three measures of sleep: duration, irregularity, and timing. All measures were derived by processing raw accelerometer data in CWA format (Continuous Wave Accelerometer), a proprietary binary format developed by Axivity. The total size of the accelerometry dataset was approximately 25TB. CWA files were processed with the Open Source R package GGIR (Version 2·3-0).^18^ This package allows for the estimation of the sleep period time window from accelerometer data by detecting periods of non-movement using the HDCZA algorithm. A detailed description of the algorithm used can be found in ^19^. To account for individual variations in calibration between accelerometers, we used the autocalibration feature of the GGIR package to calibrate measurements from individual accelerometers. Sleep timing was defined as the start and end of the longest non-movement period, while sleep duration was the length of this period. Sleep irregularity was measured as the sum of the standard deviation of the seven-day sleeping times and sleeping durations. By default, the GGIR package uses a 12:00pm sleep onset time as the pivot between ‘late’ (before midday) and ‘early’ (after midday) sleep onset. Participants were excluded for low-quality accelerometry data if: 1) they had fewer than three full nights’ worth of recordings; or 2) on included days, the participant had fewer than 16 hours recorded within the 24-maximum possible.

Processing the accelerometer data is very resource-intensive, requiring high-performance computing facilities. It took an average of 30 minutes to process data from each individual and a minimum of four GB of memory. Therefore, running the process for the whole dataset would take nearly six years on a single CPU machine. Hence, we employed Kubernetes clusters on the Google Cloud Platform to generate the outputs within a reasonable timeframe. Following initial pre-processing, post-processing was run to calculate the statistical results.

### Measurement of CVD outcomes

CVD incidence was defined as the earliest time a CVD event occurred. A CVD event was defined as myocardial infarction, heart failure, chronic ischaemic heart disease, stroke, and transient ischaemic attack. A list of outcome codes used can be found in *Supplementary Material 1*.

Participants diagnosed with CVD either before or during the period of accelerometer data collection were excluded from the analysis, along with individuals who did not have complete data for age, sex, smoking status, cholesterol levels, or hypertension, and those with diagnosed insomnia or sleep apnoea.

### Statistical Analysis

Cox proportional hazards (CPH) survival models were used to estimate hazard ratios (HRs) and 95% confidence intervals (CIs) for the association between sleep timing and risk of CVD. The follow-up time was calculated as the period from the end of accelerometer measurements to the first occurrence of CVD or the last UKB data release (2020-09-30). We also computed the mean and standard deviation for continuous measures and percentage for categorical variables.

All analyses were initially adjusted for age at recruitment and sex. A CPH model (Adjusted Model 1) was created that was additionally adjusted for sleep duration and sleep irregularity. A fully adjusted CPH model (Adjusted Model 2) was also constructed, further including other established, significant CVD risk factors: smoking status, body mass index (BMI), any diabetes, hypertension, chronotype, Townsend Deprivation Index, blood pressure, smoking status, high-density lipoprotein cholesterol, and total cholesterol.

The proportional hazards assumption was assessed for each covariate by correlating the corresponding set of scaled Schoenfeld residuals with time and testing for independence between residuals and time. A global test was also performed for the model as a whole, and no analysed variables violated the assumption of independence.

To ensure that the results were robust, we also conducted several sensitivity analyses. CPH models were constructed in subgroups stratified by age, sex, obesity, HDL cholesterol, hypertension, any diabetes, or hypotension. We also constructed models excluding participants diagnosed with CVD in the 12 to 18 months following accelerometer data collection. Results of sensitivity analyses can be found in *Supplementary Material 2, Tables 1-17*.

All statistical analyses were performed using Python version 3·7. CPH models were constructed using Python and the lifelines package. A two-sided *p*<0·05 was considered statistically significant.

## Results

### Population characteristics

Following exclusion of accelerometry participants with low-quality accelerometer data or pre-existing CVD, insomnia, or sleep apnoea, the study was conducted with 88,026 UKB participants. This included 51,214 (57·9%) females and 36,812 (41·6%) males, aged 43-79 during accelerometer data collection (mean 61·43 ± 7·8; Table 1).

**Table 1:**
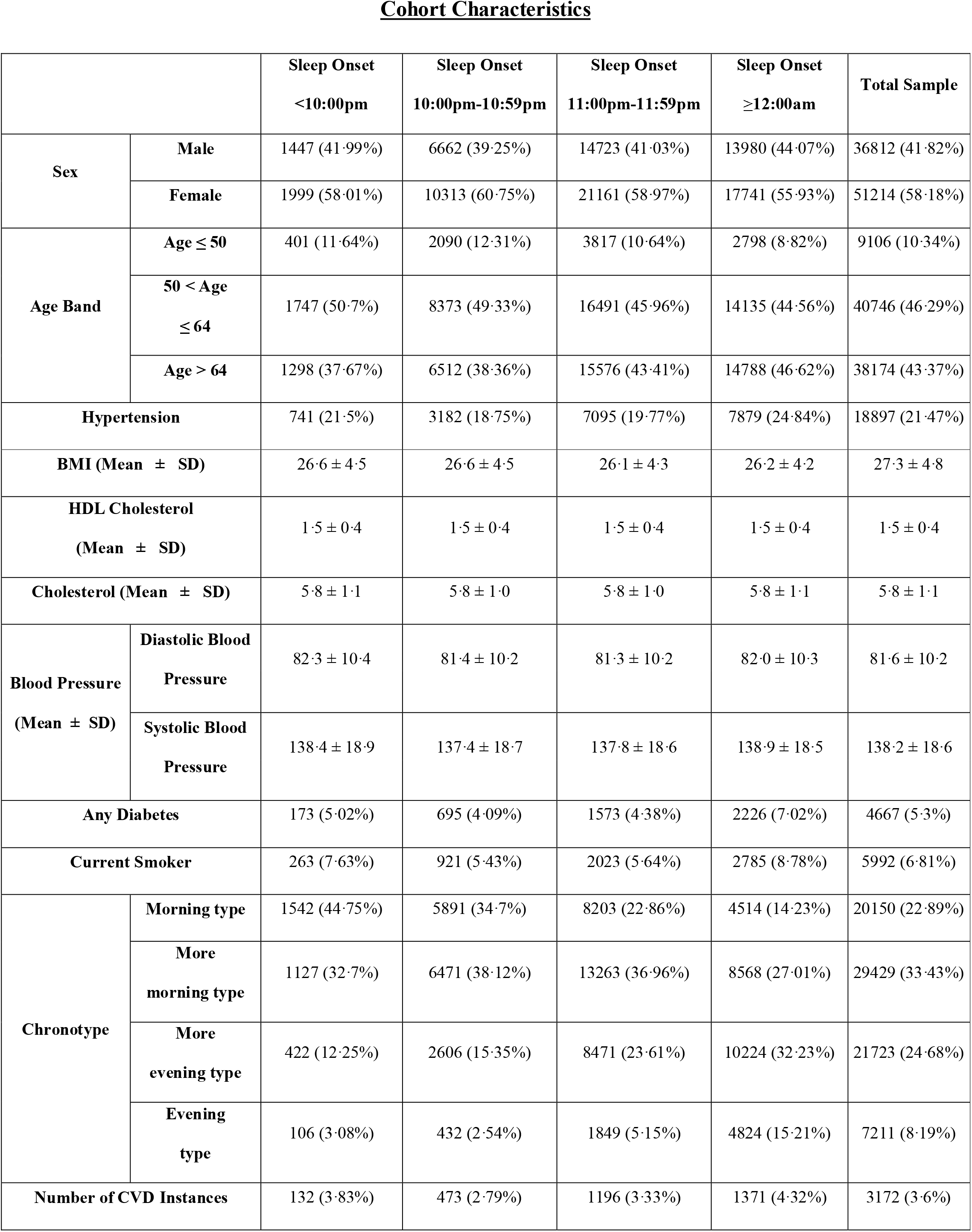
Characteristics of the cohort.

|  |  | Sleep Onset<br><10:00pm | Sleep Onset<br>10:00pm-10:59pm | Sleep Onset<br>11:00pm-11:59pm | Sleep Onset<br>≥12:00am | Total Sample |
| --- | --- | --- | --- | --- | --- | --- |
| Sex | Male | 1447 (41.99%) | 6662 (39.25%) | 14723 (41.03%) | 13980 (44.07%) | 36812 (41.82%) |
|  | Female | 1999 (58.01%) | 10313 (60.75%) | 21161 (58.97%) | 17741 (55.93%) | 51214 (58.18%) |
| Age Band | Age ≤ 50 | 401 (11.64%) | 2090 (12.31%) | 3817 (10.64%) | 2798 (8.82%) | 9106 (10.34%) |
|  | 50 < Age<br>≤ 64 | 1747 (50.7%) | 8373 (49.33%) | 16491 (45.96%) | 14135 (44.56%) | 40746 (46.29%) |
|  | Age > 64 | 1298 (37.67%) | 6512 (38.36%) | 15576 (43.41%) | 14788 (46.62%) | 38174 (43.37%) |
| Hypertension |  | 741 (21.5%) | 3182 (18.75%) | 7095 (19.77%) | 7879 (24.84%) | 18897 (21.47%) |
| BMI (Mean ± SD) |  | 26.6 ± 4.5 | 26.6 ± 4.5 | 26.1 ± 4.3 | 26.2 ± 4.2 | 27.3 ± 4.8 |
| HDL Cholesterol<br>(Mean ± SD) |  | 1.5 ± 0.4 | 1.5 ± 0.4 | 1.5 ± 0.4 | 1.5 ± 0.4 | 1.5 ± 0.4 |
| Cholesterol (Mean ± SD) |  | 5.8 ± 1.1 | 5.8 ± 1.0 | 5.8 ± 1.0 | 5.8 ± 1.1 | 5.8 ± 1.1 |
| Blood Pressure<br>(Mean ± SD) | Diastolic Blood<br>Pressure | 82.3 ± 10.4 | 81.4 ± 10.2 | 81.3 ± 10.2 | 82.0 ± 10.3 | 81.6 ± 10.2 |
|  | Systolic Blood<br>Pressure | 138.4 ± 18.9 | 137.4 ± 18.7 | 137.8 ± 18.6 | 138.9 ± 18.5 | 138.2 ± 18.6 |
| Any Diabetes |  | 173 (5.02%) | 695 (4.09%) | 1573 (4.38%) | 2226 (7.02%) | 4667 (5.3%) |
| Current Smoker |  | 263 (7.63%) | 921 (5.43%) | 2023 (5.64%) | 2785 (8.78%) | 5992 (6.81%) |
| Chronotype | Morning type | 1542 (44.75%) | 5891 (34.7%) | 8203 (22.86%) | 4514 (14.23%) | 20150 (22.89%) |
|  | More<br>morning type | 1127 (32.7%) | 6471 (38.12%) | 13263 (36.96%) | 8568 (27.01%) | 29429 (33.43%) |
|  | More<br>evening type | 422 (12.25%) | 2606 (15.35%) | 8471 (23.61%) | 10224 (32.23%) | 21723 (24.68%) |
|  | Evening<br>type | 106 (3.08%) | 432 (2.54%) | 1849 (5.15%) | 4824 (15.21%) | 7211 (8.19%) |
| Number of CVD Instances |  | 132 (3.83%) | 473 (2.79%) | 1196 (3.33%) | 1371 (4.32%) | 3172 (3.6%) |

During follow-up (mean 5·7 ±0·49 years), there were 3172 participants (3·58%) who developed CVD, of which: 1371 (43%) had a sleep onset time (SOT) after midnight; 1196 (38%) had an SOT between 11:00pm and 11:59pm; 473 (15%) had an SOT between 10:00pm and 10:59pm; and 132 (4·2%) had an SOT before 10pm (Table 1). Additionally, Kaplan-Meier curves for each SOT category showed the rate of CVD to be the highest in participants with SOTs ≥12am and the lowest when SOT is between 10:00pm to 10:59pm (Table 1 and Figure 2).

**Figure 1:**
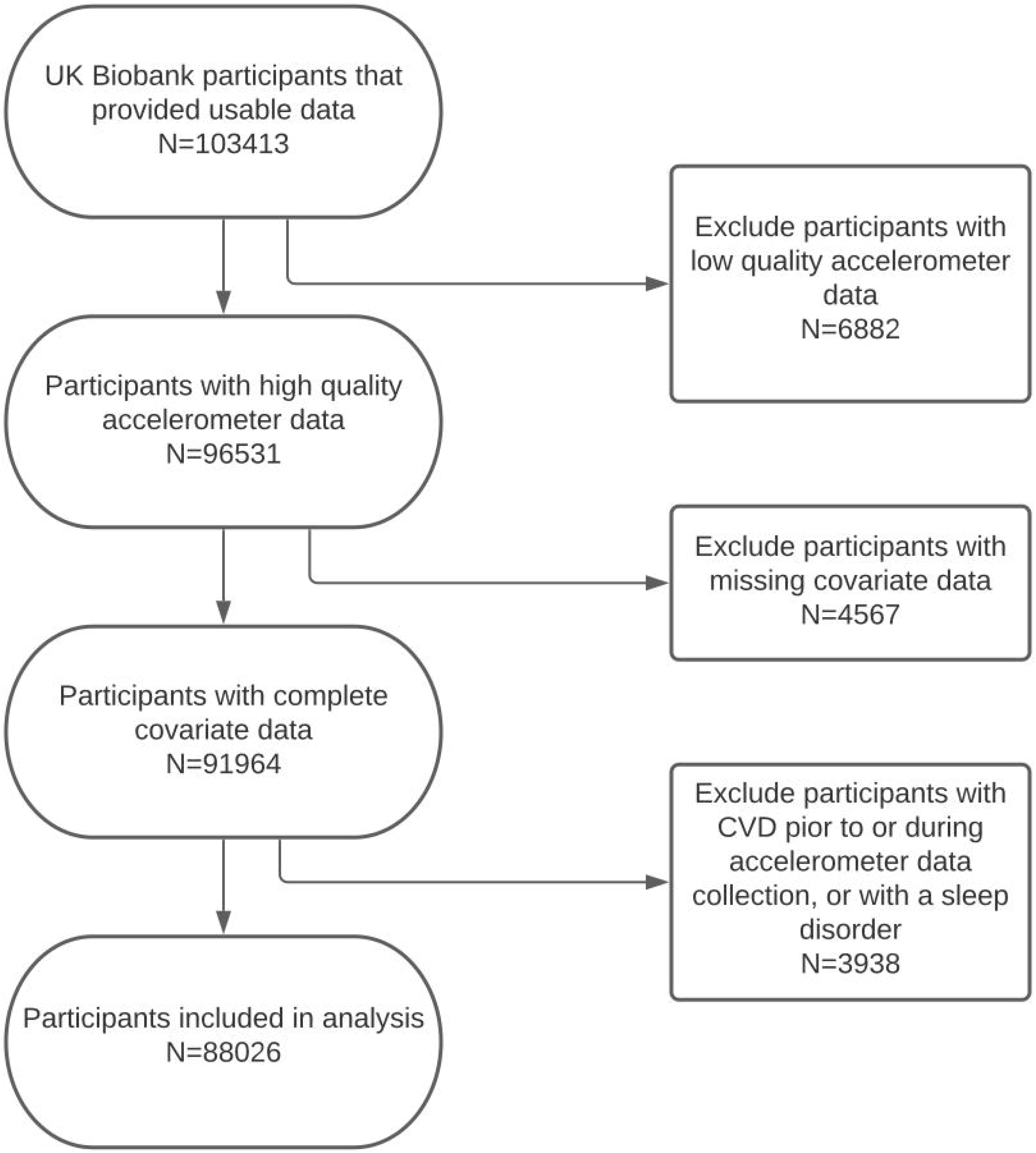
Flow of participants through the study.

**Figure 2:**
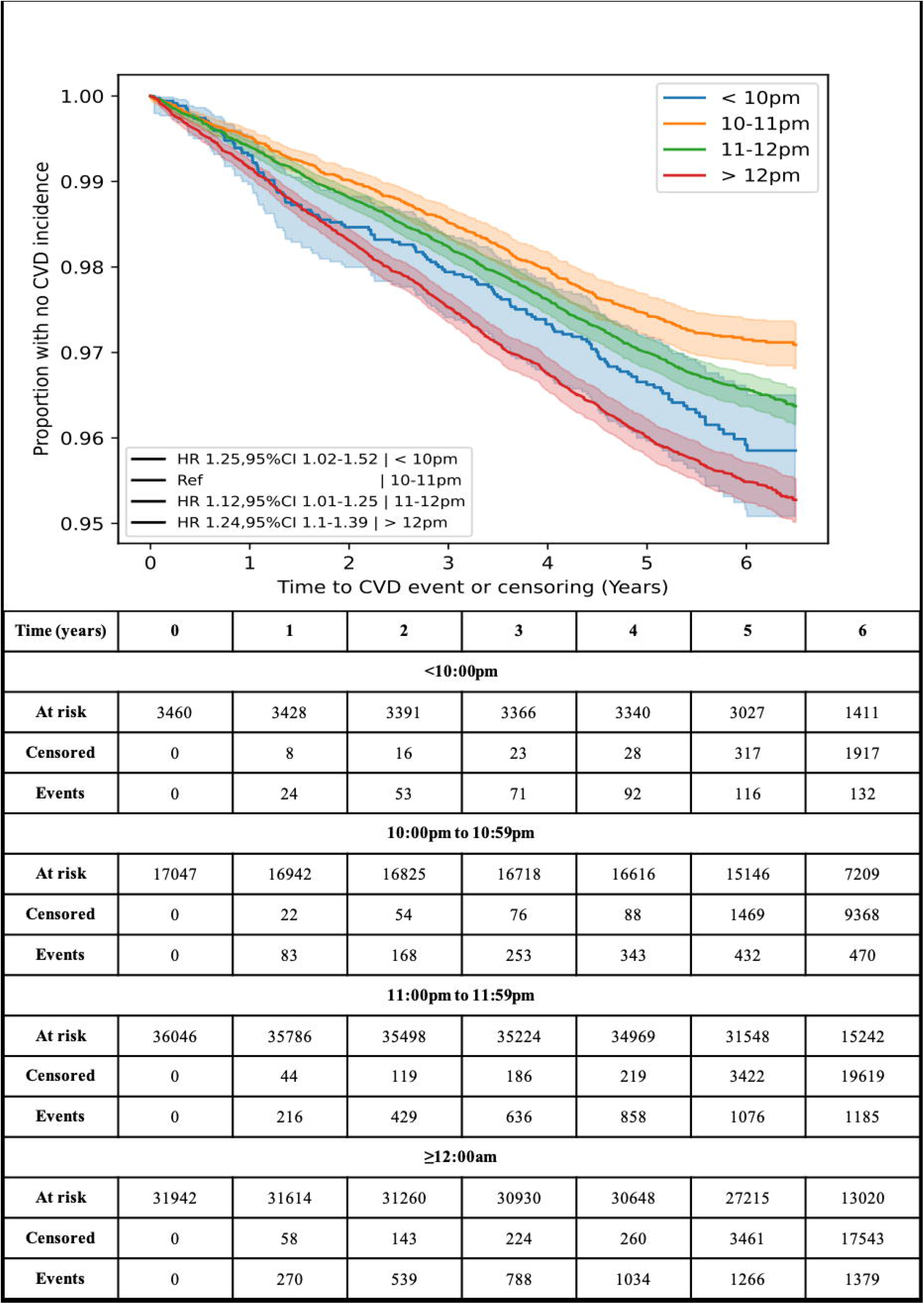
Kaplan-Meier curves for cardiovascular disease incidence against time to occurrence, split by sleep onset time (SOT).

Participants with an SOT after midnight (n=31,946) were more likely to be a current smoker and have a history of diabetes or hypertension than those with SOTs from 11:00pm to 11:59pm (n=36,042), 10:00pm to 10:59pm (n=17,040), and before 10:00pm (n=3460).

### Assessment of SOTs, Sleep Irregularity, and Sleep Duration

There was a U–shaped relationship between increased incidence of CVD and SOT, with a higher rate of incidence for participants with late or very early SOTs (Figure 3). There were 3·82, 2·78, 3·32, and 4·29 CVD incidences per 100 people in SOT categories of <10:00pm, 10:00pm-10:59pm, 11:00pm-11:59pm, and ≥12:00am, respectively.

**Figure 3:**
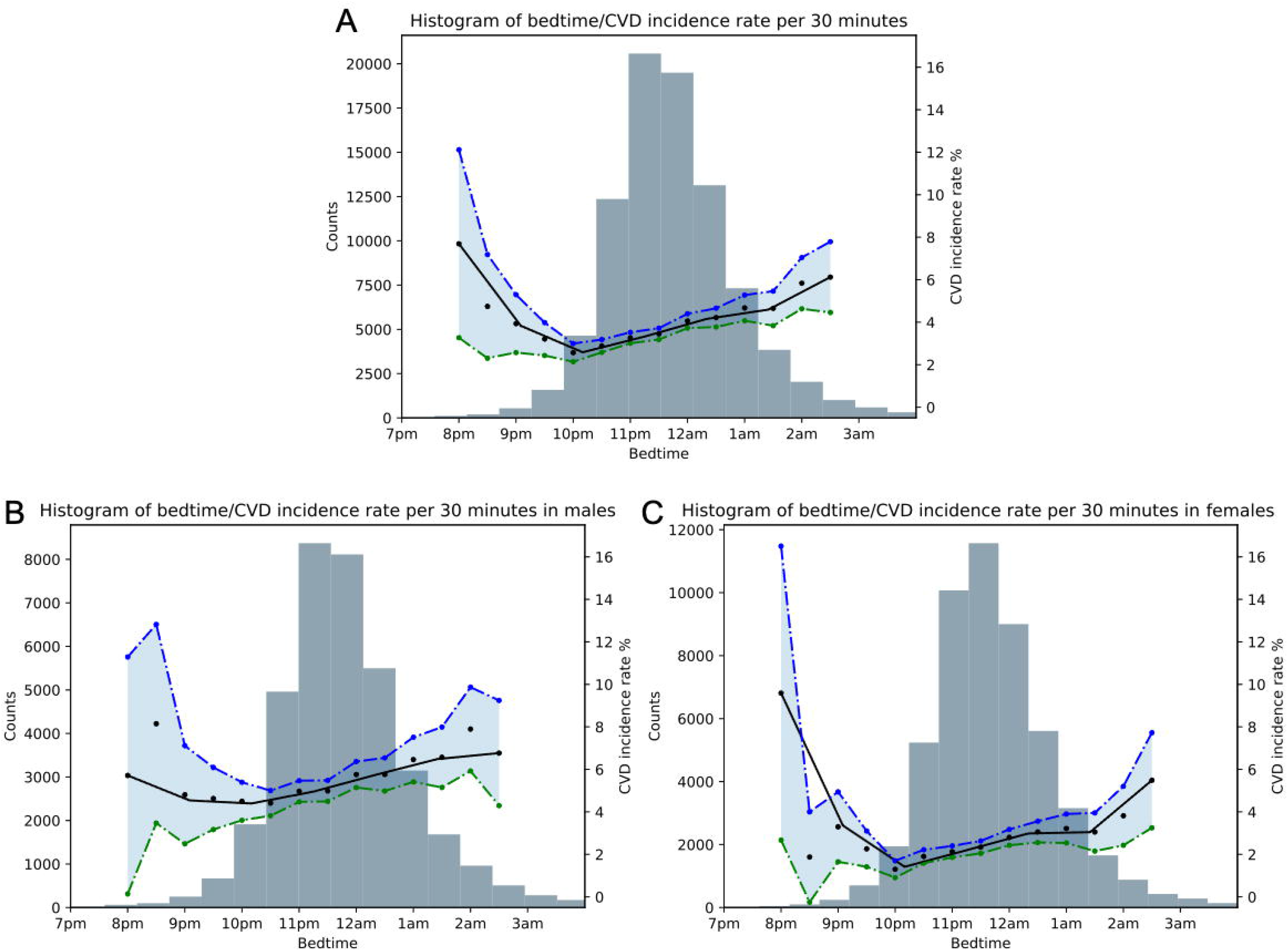
Histogram showing the relationship between sleep onset time (SOT) and CVD incidence rate for: A) the overall sample, B) males and C) females. The black line indicates the mean CVD incidence rate per 30-minute increments. Blue and green lines indicate 95% confidence intervals.

In the baseline analysis, the group with SOTs between 10:00pm and 10:59pm had the lowest incidence rate of CVD and was chosen as the reference group. The HRs obtained were 1·35 (95% CI, 1·12–1·64; *p*<0·005), 1·11 (95% CI, 1·00–1·24; *p*=0·05), and 1·38 (95% CI, 1·24–1·53; *p*<0·005) for SOT categories ≥12am, 11pm-11:59pm, and <10pm, respectively.

After adjusting for sleep duration and sleep irregularity (Adjusted Model 1), the HRs obtained were 1·29 (95% CI, 1·06–1·57; *p*=0·01), 1·09 (95% CI, 0·98–1·22; *p*=0·1), and 1·25 (95% CI, 1·12– 1·39; *p*<0·005) for SOT categories ≥12:00am, 11:00pm-11:59pm, and <10:00pm, respectively (Table 2). Moreover, the HR for sleep duration was 0·93 (0·90–0·96), indicating that shorter sleep duration is associated with increased CVD risk. Similarly, the HR for sleep irregularity was 1·04 (1·02-1·07), mirroring the findings of prior research that irregular sleep is associated with an increased risk of CVD.

**Table 2:** Cox proportional hazard models hazard ratios for sleep onset time (SOT). The Base Model includes age and sex as covariates. Adjusted Model 1 is adjusted for Base Model features, sleep duration, and sleep irregularity. Adjusted Model 2 is adjusted for Adjusted Model 1 features, BMI, pre-existing diabetes, pre-existing hypertension, chronotype, Townsend Deprivation Index, blood pressure, smoking status, high-density lipoprotein cholesterol, and total cholesterol. CI = confidence interval.

|  | Base Model |  | Adjusted Model 1 |  | Fully Adjusted Model 2 |  |
| --- | --- | --- | --- | --- | --- | --- |
|  | Hazard Ratio (95% CI) | *p*-value | Hazard Ratio (95% CI) | *p*-value | Hazard Ratio (95% CI) | *p*-value |
| **≥12:00am** | 1.35 (1.12-1.64) | < 0.005 | 1.29 (1.06-1.57) | 0.01 | 1.25 (1.02-1.52) | 0.03 |
| **11:00pm-11:59pm** | 1.11 (1.00-1.24) | 0.05 | 1.09 (0.98-1.22) | 0.1 | 1.12 (1.01-1.25) | 0.04 |
| **10:00pm-10:59pm** | 1 (ref) | .. | 1 (ref) | .. | 1 (ref) | .. |
| **<10:00pm** | 1.38 (1.24-1.53) | < 0.005 | 1.25 (1.12-1.39) | < 0.005 | 1.24 (1.10-1.39) | < 0.005 |
| **Sleep Duration** | .. | .. | 0.93 (0.90-0.96) | < 0.005 | 0.97 (0.94-1.00) | 0.04 |
| **Sleep Irregularity** | .. | .. | 1.04 (1.02-1.07) | < 0.005 | 1.02 (0.99-1.05) | 0.12 |

After adjustment for established CVD risk factors (Adjusted Model 2), the HRs decreased slightly but remained equally statistically significant. The fully adjusted HRs for the different SOT categories were 1·25 (95% CI, 1·02–1·52; *p*=0·03) in the ≥12:00am, 1·12 (95% CI, 1·01–1·25; *p*=0·04) in the 11:00pm-11:59pm group, and 1·24 (95% CI, 1·10–1·39; *p*<0·005) in the <10:00pm group. Similarly, the HRs for sleep duration were 0·97 (95% CI, 0·94–1·00; *p*=0·04) in the fully adjusted model, and 1·02 (95% CI, 0·99–1·05; *p*=0·12) for sleep irregularity (Table 2).

However, a sensitivity analysis where models were constructed for males and females separately revealed that SOTs ≥12:00am (HR=1·63; 95% CI, 1·20–2·21; *p*<0·005) and <10:00pm (HR=1·34; 95% CI, 1·11–1·61; *p*<0·005) were significantly associated with CVD risk in females, while only SOTs <10:00pm (HR=1·17; 95% CI, 1·01–1·35; *p*=0·03) were significantly associated with increased CVD risk in males, after controlling for all fully Adjusted Model 2 features (Table 3; full details in *Supplementary Tables 11-12*).

**Table 3:** Sensitivity analysis for sleep onset time (SOT) in males and females using the fully Adjusted Model 2 covariates. CI = confidence interval.

|  | Male (N=36,812) |  |  |  | Female (N=51,214) |  |  |  |
| --- | --- | --- | --- | --- | --- | --- | --- | --- |
| Fully<br>Adjusted<br>Model 2 | ≥12:00am | 11:00pm-11:59pm | 10:00pm-10:59pm | <10:00pm | ≥12:00am | 11:00pm-11:59pm | 10:00pm-10:59pm | <10:00pm |
| Hazard Ratio<br>(95% CI) | 1·05<br>(0·81-1·35) | 1·08<br>(0·94-1·24) | 1 (ref) | 1·17<br>(1·01-1·35) | 1·63<br>(1·20-2·21) | 1·18<br>(0·99-1·40) | 1 (ref) | 1·34<br>(1·11-1·61) |
| *p*-value | 0·73 | 0·27 | .. | 0·03 | < 0·005 | 0·07 | .. | < 0·005 |

## Discussion

To the best of our knowledge, this is the most extensive study to date to investigate objectively assessed sleep parameters through the use of accelerometry. We have demonstrated a clear association between timing of sleep and CVD risk, particularly for women. While later SOTs were associated with an increased incidence of CVD, the relationship was U-shaped, agreeing with the idea that the optimum SOT falls within a specific range of the diurnal cycle and that deviations from this range, either too early or too late, are problematic. The highest incidence of CVD was found in participants with SOTs after midnight, with the incidence of CVD falling with earlier SOTs, before rising again with SOTs <10pm. Our findings conform with one previous study where late bedtimes (>11:00pm) on weekdays were associated with an increased risk of congestive heart failure.^15^

The relationship between sleep timing and CVD is understudied. While sleep quality can be divided into sleep timing, sleep length, and sleep irregularity, our investigation focused on sleep timing as an independent variable owing to its prior lack of study. The association between sleep outside of 10pm-11pm and CVD persisted even after adjustment for sleep duration and sleep irregularity. This suggests that the timing of sleep can affect CVD risk independently of other features that contribute to ‘poor’ sleep quality. To ensure that present-but-undiagnosed CVD was not causing disturbed sleep timing and biasing the results, we carried out sensitivity analyses where either the first 12- or 18-months following accelerometer data collection were excluded (*Supplementary Table 16-17*). However, the overall associations between sleep onset timing and CVD risk persisted.

Further sensitivity analyses concerning age categories (*Supplementary Table 13-15*) and pre-existing conditions (*Supplementary Table 1-9*) similarly did not impact the overall association, however, sex constitutes a significant exception (Table 3; *Supplementary Tables 11-12*). Constructing separate models in males and females revealed a dimorphic pattern whereby the associations between sleep onset timing and CVD were significantly more robust with greater weights for females, than males. In men, only <10pm remained significant but with small associated risk, potentially suggesting that confounders influencing earlier SOT were unaccounted for, such as other pre-existing conditions, rather than a true association between sleep timing and CVD.

Differences in cardiovascular risk between women and men are long-established, with CVD classically described as a disease affecting elderly western males, in part due to women typically carrying a degree of cardioprotection. Post-menopause, however, rates of CVD in women rise to meet or even surpass male counterparts, likely resulting from decreased oestrogen.^20^ Moreover, the cellular composition of the heart differs between the sexes, with variance in cardiac electrophysiology, lipid membrane composition, mitochondria, and myofilament composition.^12^ Our findings, however, suggest an increased risk of CVD for women in the presence of circadian rhythm disruption from sleep timing, independent of other factors. The reasons behind this are unclear. Other preliminary research indicates significant differences in circadian endocrine rhythms between the sexes.^21^ Several anatomical and functional cardiovascular differences have been identified between males and females in both animal and human studies.^12^ However, the cellular mechanisms behind these are still unclear. A recent study also supports the idea of a sex-specific impact of suboptimal sleep in CVD, finding that disrupted sleep may have a more significant impact on CVD mortality risk in women than men.^22^

There is substantial evidence that sleep is related to other CVD risk factors, such as obesity and hypertension, with later bedtime in particular correlated with higher BMI.^23,24^ Timing of sleep has also been demonstrated to be negatively associated with glycaemic control in patients with type 2 diabetes mellitus.^25^ Jansen et al. found that Mexican adolescents with a weekday bedtime later than 11pm had a 1·87 times higher risk of developing elevated BP than the participants with a bedtime between 9pm and 10pm, after accounting for sleep-related confounders.^26^ In our study, however, the association between timing of sleep and CVD incidence remained robust even after additional adjustment for important CVD risk factors, such as hypertension, diabetes, BMI, and smoking.

Notably, two of the classically described contributors to CVD in the event of circadian rhythm disturbance are raised blood pressure and obesity. However, these potential confounders are also accounted for in this study. The pathophysiological process behind any increased risk is, therefore, unclear. Potential contributors could be the disruption of endocrine processes aligned with the circadian rhythm.^27^ Such disruption would increase CVD risk through physiological processes independent of hypercholesterolaemia, hypertension, and diabetes. Potential processes could include endothelial dysfunction, prothrombotic states, or platelet dysfunction.^13^

Although the findings of this paper do not show causality, they mandate further research into sleep timing as an independent cardiac risk factor, particularly for women. Sleep timing would be an attractive target for interventions to reduce CVD risk owing to its minimal cost and invasiveness. This intervention could take the form of public health guidance, structured intervention programs, or technology-based solutions such as smartphone apps. Although it was not possible to focus on them in the modelling, sleep timing as a CVD risk factor would have considerable implications on night-shift workers, the organisations of their schedules, and the ethics of such work.

Importantly, this study demonstrates the value of using objectively collected ‘big data’ to assess sleep parameters as an alternative to subjective sleep diaries or journals, which are often significantly inaccurate.^28^ Wearable devices have the potential to improve real-time CVD risk surveillance through the passive collection of accelerometer data, which simultaneously can enable the elimination of the recall bias associated with retrospective questionnaires or interviews.

The major strength of this study is the UKB dataset: an unparalleled, longitudinal dataset encompassing non-traditional health data, such as accelerometer data, alongside genomics, prospective health record integration, and additional blood markers. Notably, most prior sleep investigations have used subjective reports of bedtime and sleep parameters, while our investigation is one of the largest studies employing objectively measured sleep parameters. Our use of objectively derived accelerometer data avoids the error due to recall bias that results from assessment using questionnaires.

However, the UKB cohort is predominantly White British and has an overrepresentation of individuals from higher socioeconomic backgrounds, resulting in its ‘healthier and wealthier’ phenomenon.^29^ This could mean that the presented findings may not generalise well to other populations, and further research in large samples more representative of the global population is required. Moreover, while the UKB has some records of night-shift work, only ∼5% of the cohort report ever engaging with it. This, in addition to the fact that occupational data was collected 3-10 years prior to accelerometry and includes no historical data on shift-working frequency, meant we could not reliably assess this high-risk demographic in isolation.

Although multiple studies validate the accuracy of sleep measurement via actigraphy, this method has some limitations. We found that the analysis of the accelerometer data did not perform very well if the participant had abnormal activity during sleep. Additionally, the HDCZA algorithm used to determine sleep timing, duration, and irregularity is imperfect with a c-statistic of 0·83 to detect the sleep period time window compared to polysomnography in those without sleep disorders. Last, a seven-day measurement is not necessarily the best representation of habitual sleeping, albeit standard in the field. While we accounted for the majority of traditional risk factors as potential confounders, limitations in the data meant that we could not include family history in our modelling in a meaningful way.

Overall, our findings suggest an independent relationship between sleep onset timing and risk of developing cardiovascular disease, particularly in women. As dimorphic patterns were seen between the sexes, we recommend further research into the potential underlying mechanisms and prospective studies from other geographies. Nevertheless, sleep timing represents an understudied and potentially novel risk factor that may be a valuable target for public health guidance in primary prevention of CVD. Our results also demonstrate the efficacy and convenience of passive, accelerometer-derived big data as a predictor of health outcomes.

## Data Availability

The UKB dataset was obtained from the UKB (application number 55668). Data cannot be shared publicly owing to the violation of patient privacy and the absence of informed consent for data sharing.

## Authors’ contributions

SN, ABR, ACC, BDO, MA, DM and DP drafted and reviewed the manuscript. All authors approved the manuscript before submission. SM, ABR, ACC, BDO, MA, DM and DP analysed and interpreted the data.

## References

1 Vos T, Lim SS, Abbafati C, et al. Global burden of 369 diseases and injuries in 204 countries and territories, 1990–2019: a systematic analysis for the Global Burden of Disease Study 2019. The Lancet 2020; 396: 1204–22.

2 Brown JC, Gerhardt TE, Kwon E. Risk Factors For Coronary Artery Disease. In: StatPearls. Treasure Island (FL): StatPearls Publishing, 2021. http://www.ncbi.nlm.nih.gov/books/NBK554410/ (accessed May 27, 2021).

3 Burgess E, Hassmén P, Welvaert M, Pumpa KL. Behavioural treatment strategies improve adherence to lifestyle intervention programmes in adults with obesity: a systematic review and meta-analysis. Clin Obes 2017; 7: 105–14.

4 Michie S, van Stralen MM, West R. The behaviour change wheel: A new method for characterising and designing behaviour change interventions. Implement Sci 2011; 6: 42.

5 Scheer FAJL, Hilton MF, Mantzoros CS, Shea SA. Adverse metabolic and cardiovascular consequences of circadian misalignment. Proc Natl Acad Sci 2009; 106: 4453–8.

6 Portaluppi F, Tiseo R, Smolensky MH, Hermida RC, Ayala DE, Fabbian F. Circadian rhythms and cardiovascular health. Sleep Med Rev 2012; 16: 151–66.

7 Morris CJ, Purvis TE, Hu K, Scheer Fajl. Circadian misalignment increases cardiovascular disease risk factors in humans. Proc Natl Acad Sci 2016; 113: E1402–11.

8 Atkinson G, Davenne D. Relationships between sleep, physical activity and human health. Physiol Behav 2007; 90: 229–35.

9 Zhao R, Li D, Zuo P, et al. Influences of Age, Gender, and Circadian Rhythm on Deceleration Capacity in Subjects without Evident Heart Diseases. Ann Noninvasive Electrocardiol Off J Int Soc Holter Noninvasive Electrocardiol Inc 2014; 20: 158–66.

10 Yang L, Chu Y, Wang L, et al. Overexpression of CRY1 protects against the development of atherosclerosis via the TLR/NF-κB pathway. Int Immunopharmacol 2015; 28: 525–30.

11 Pan X, Jiang X-C, Hussain MM. Impaired Cholesterol Metabolism and Enhanced Atherosclerosis in Clock Mutant Mice. Circulation 2013; 128: 1758–69.

12 Glen Pyle W, Martino TA. Circadian rhythms influence cardiovascular disease differently in males and females: role of sex and gender. Curr Opin Physiol 2018; 5: 30–7.

13 Crnko S, Du Pré BC, Sluijter JPG, Van Laake LW. Circadian rhythms and the molecular clock in cardiovascular biology and disease. Nat Rev Cardiol 2019; 16: 437–47.

14 Huang T, Mariani S, Redline S. Sleep Irregularity and Risk of Cardiovascular Events: The Multi-Ethnic Study of Atherosclerosis. J Am Coll Cardiol 2020; 75: 991–9.

15 Yan B, Li R, Li J, et al. Sleep Timing May Predict Congestive Heart Failure: A Community□Based Cohort Study. J Am Heart Assoc 2021; 10: e018385.

16 Jørgensen JT, Karlsen S, Stayner L, Hansen J, Andersen ZJ. Shift work and overall and cause-specific mortality in the Danish nurse cohort. Scand J Work Environ Health 2017; 43: 117–26.

17 Doherty A, Jackson D, Hammerla N, et al. Large Scale Population Assessment of Physical Activity Using Wrist Worn Accelerometers: The UK Biobank Study. PLOS ONE 2017; 12: e0169649.

18 Migueles JH, Rowlands AV, Huber F, Sabia S, Hees VT van. GGIR: A Research Community–Driven Open Source R Package for Generating Physical Activity and Sleep Outcomes From Multi-Day Raw Accelerometer Data. J Meas Phys Behav 2019; 2: 188–96.

19 van Hees VT, Sabia S, Jones SE, et al. Estimating sleep parameters using an accelerometer without sleep diary. Sci Rep 2018; 8: 12975.

20 Mozaffarian D, Benjamin EJ, Go AS, et al. Executive Summary: Heart Disease and Stroke Statistics—2016 Update. Circulation 2016; 133: 447–54.

21 Nicolaides NC, Chrousos GP. Sex differences in circadian endocrine rhythms: Clinical implications. Eur J Neurosci 2020; 52: 2575–85.

22 Shahrbabaki SS, Linz D, Hartmann S, Redline S, Baumert M. Sleep arousal burden is associated with long-term all-cause and cardiovascular mortality in 8001 community-dwelling older men and women. Eur Heart J 2021; published online April 20. DOI:10.1093/eurheartj/ehab151.

23 Asarnow LD, McGlinchey E, Harvey AG. Evidence for a Possible Link between Bedtime and Change in Body Mass Index. Sleep 2015; 38: 1523–7.

24 Chaput J-P, Dutil C, Featherstone R, et al. Sleep timing, sleep consistency, and health in adults: a systematic review1. Appl Physiol Nutr Metab 2020; published online Oct 15. DOI:10.1139/apnm-2020-0032.

25 Reutrakul S, Siwasaranond N, Nimitphong H, et al. Relationships among sleep timing, sleep duration and glycemic control in Type 2 diabetes in Thailand. Chronobiol Int 2015; 32: 1469–76.

26 Jansen EC, Dunietz GL, Matos-Moreno A, Solano M, Lazcano-Ponce E, Sánchez-Zamorano LM. Bedtimes and Blood Pressure: A Prospective Cohort Study of Mexican Adolescents. Am J Hypertens 2020; 33: 269–77.

27 Bedrosian TA, Fonken LK, Nelson RJ. Endocrine Effects of Circadian Disruption. Annu Rev Physiol 2016; 78: 109–31.

28 Kong N, Choi J, Seo WS. Evaluation of Sleep Problems or Disorders Using Sleep Questionnaires. Chronobiol Med 2019; 1: 144–8.

29 Fry A, Littlejohns TJ, Sudlow C, et al. Comparison of Sociodemographic and Health-Related Characteristics of UK Biobank Participants With Those of the General Population. Am J Epidemiol 2017; 186: 1026–34.

