## Supplementary Material for "Accelerometer-derived sleep onset timing and cardiovascular disease incidence: a UK Biobank cohort study"

### **Supplementary Material 1 - Codes used to identify CVD events from UK Biobank**

Myocardial infarction is identified by UK Biobank field IDs (42000, 131298, 131300, 131302, and 131304) and ICD10 codes (I21*-I25*).

Heart failure is identified by UK Biobank field ID (131354) and ICD10 codes (I50*, I11·0, I13·0, and I13·2).

Chronic ischaemic heart diseases are identified by UK Biobank field IDs (131296 and 131306) and ICD10 codes (I20*).

Stroke is identified by UK Biobank field IDs (42006, 131360, 131362, 131366, and 131368) and ICD10 codes (G46*, I60*-I64*, I67·81, and I67·82).

Transient Ischaemic Attack is identified by UK Biobank field ID (131056) and ICD10 codes (G45*).

42000 - Algorithmically-defined outcomes (Myocardial infarction)

131298 - First occurrences (Acute myocardial infarction)

131300 - First occurrences (Subsequent myocardial infarction)

131302 - First occurrences (Complications after myocardial infarction)

131304 - First occurrences (Acute ischaemic heart disease)

131354 - First occurrences (Heart failure)

131296 - First Occurrences (Angina Pectoris)

131306 - First occurrences (Chronic ischaemic heart disease)

42006 - Algorithmically-defined outcomes (Stroke)

131360 - First occurrences (Subarachnoid haemorrhage)

131362 - First occurrences (Intracerebral haemorrhage)

131366 - First occurrences (Cerebral infarction)

131368 - First occurrences (Stroke unspecified)

131056 - First Occurrences (Transient Ischaemic Attack)

### **Supplementary Material 2 - Sensitivity analyses**

|  | **Base Model** | | **Adjusted Model 1** | | **Fully Adjusted Model 2** | |
| --- | --- | --- | --- | --- | --- | --- |
| **Sleep Onset Time** | **Hazard Ratio (95% CI)** | ***p*-value** | **Hazard Ratio (95% CI)** | ***p*-value** | **Hazard Ratio (95% CI)** | ***p*-value** |
| **≥12:00am** | 1·18 (0·91-1·52) | 0·21 | 1·14 (0·88-1·48) | 0·31 | 1·14 (0·88-1·48) | 0·32 |
| **11:00pm-11:59pm** | 1·14 (0·99-1·31) | 0·07 | 1·13 (0·98-1·30) | 0·1 | 1·15 (1·00-1·32) | 0·06 |
| **10:00pm-10:59pm** | 1 (ref) | .. | 1 (ref) | .. | 1 (ref) | .. |
| **<10:00pm** | 1·25 (1·09-1·43) | < 0·005 | 1·16 (1·01-1·34) | 0·04 | 1·22 (1·05-1·42) | 0·01 |
| **Sleep Duration** | .. | .. | 0·95 (0·91-0·99) | 0·02 | 0·96 (0·92-1·00) | 0·08 |
| **Sleep Irregularity** | .. | .. | 1·02 (0·99-1·06) | 0·15 | 1·02 (0·99-1·05) | 0·26 |

**Supplementary Table 1:** Results of sensitivity analysis in participants with hypertension (N=18897).

|  | **Base Model** | | **Adjusted Model 1** | | **Fully Adjusted Model 2** | |
| --- | --- | --- | --- | --- | --- | --- |
| **Sleep Onset Time** | **Hazard Ratio (95% CI)** | ***p*-value** | **Hazard Ratio (95% CI)** | ***p*-value** | **Hazard Ratio (95% CI)** | ***p*-value** |
| **≥12:00am** | 1·46 (1·09-1·95) | 0·01 | 1·42 (1·06-1·91) | 0·02 | 1·41 (1·05-1·90) | 0·02 |
| **11:00pm-11:59pm** | 1·09 (0·92-1·28) | 0·31 | 1·08 (0·91-1·27) | 0·38 | 1·07 (0·91-1·27) | 0·39 |
| **10:00pm-10:59pm** | 1 (ref) | .. | 1 (ref) | .. | 1 (ref) | .. |
| **<10:00pm** | 1·33 (1·13-1·57) | < 0·005 | 1·26 (1·06-1·49) | 0·01 | 1·23 (1·03-1·47) | 0·02 |
| **Sleep Duration** | .. | .. | 0·96 (0·91-1·01) | 0·11 | 0·97 (0·92-1·02) | 0·2 |
| **Sleep Irregularity** | .. | .. | 1·03 (0·99-1·07) | 0·19 | 1·02 (0·98-1·06) | 0·35 |

**Supplementary Table 2:** Results of sensitivity analysis in participants without hypertension (N=75426).

|  | **Base Model** | | **Adjusted Model 1** | | **Fully Adjusted Model 2** | |
| --- | --- | --- | --- | --- | --- | --- |
| **Sleep Onset Time** | **Hazard Ratio (95% CI)** | ***p*-value** | **Hazard Ratio (95% CI)** | ***p*-value** | **Hazard Ratio (95% CI)** | ***p*-value** |
| **≥12:00am** | 1·27 (0·86-1·88) | 0·23 | 1·21 (0·81-1·80) | 0·35 | 1·18 (0·79-1·76) | 0·41 |
| **11:00pm-11:59pm** | 1·05 (0·83-1·31) | 0·71 | 1·02 (0·81-1·28) | 0·88 | 1·07 (0·85-1·35) | 0·55 |
| **10:00pm-10:59pm** | 1 (ref) | .. | 1 (ref) | .. | 1 (ref) | .. |
| **<10:00pm** | 1·33 (1·07-1·65) | 0·01 | 1·16 (0·93-1·45) | 0·19 | 1·22 (0·97-1·54) | 0·09 |
| **Sleep Duration** | .. | .. | 0·89 (0·84-0·94) | < 0·005 | 0·92 (0·87-0·98) | 0·01 |
| **Sleep Irregularity** | .. | .. | 1·03 (0·98-1·07) | 0·27 | 1·00 (0·96-1·05) | 0·97 |

**Supplementary Table 3:** Results of sensitivity analysis in participants who were obese (N= 16296).

|  | **Base Model** | | **Adjusted Model 1** | | **Fully Adjusted Model 2** | |
| --- | --- | --- | --- | --- | --- | --- |
| **Sleep Onset Time** | **Hazard Ratio (95% CI)** | ***p*-value** | **Hazard Ratio (95% CI)** | ***p*-value** | **Hazard Ratio (95% CI)** | ***p*-value** |
| **≥12:00am** | 1·36 (1·09-1·69) | 0·01 | 1·30 (1·04-1·62) | 0·02 | 1·27 (1·01-1·59) | 0·04 |
| **11:00pm-11:59pm** | 1·13 (1·00-1·27) | 0·05 | 1·12 (0·99-1·26) | 0·07 | 1·14 (1·01-1·28) | 0·04 |
| **10:00pm-10:59pm** | 1 (ref) | .. | 1 (ref) | .. | 1 (ref) | .. |
| **<10:00pm** | 1·33 (1·18-1·50) | < 0·005 | 1·25 (1·10-1·42) | < 0·005 | 1·25 (1·10-1·43) | < 0·005 |
| **Sleep Duration** | .. | .. | 0·97 (0·93-1·00) | 0·08 | 0·98 (0·95-1·02) | 0·43 |
| **Sleep Irregularity** | .. | .. | 1·04 (1·01-1·07) | < 0·005 | 1·03 (1·00-1·06) | 0·07 |

**Supplementary Table 4:** Results of sensitivity analysis in participants who were not obese (N=71719).

|  | **Base Model** | | **Adjusted Model 1** | | **Fully Adjusted Model 2** | |
| --- | --- | --- | --- | --- | --- | --- |
| **Sleep Onset Time** | **Hazard Ratio (95% CI)** | ***p*-value** | **Hazard Ratio (95% CI)** | ***p*-value** | **Hazard Ratio (95% CI)** | ***p*-value** |
| **≥12:00am** | 1·67 (1·01-2·78) | 0·05 | 1·47 (0·87-2·47) | 0·15 | 1·40 (0·83-2·35) | 0·21 |
| **11:00pm-11:59pm** | 1·16 (0·84-1·60) | 0·36 | 1·16 (0·84-1·60) | 0·37 | 1·20 (0·87-1·66) | 0·27 |
| **10:00pm-10:59pm** | 1 (ref) | .. | 1 (ref) | .. | 1 (ref) | .. |
| **<10:00pm** | 1·47 (1·09-1·99) | 0·01 | 1·30 (0·95-1·78) | 0·1 | 1·36 (0·99-1·88) | 0·06 |
| **Sleep Duration** | .. | .. | 0·93 (0·86-1·01) | 0·08 | 0·96 (0·89-1·04) | 0·3 |
| **Sleep Irregularity** | .. | .. | 1·07 (1·01-1·13) | 0·02 | 1·05 (0·99-1·12) | 0·08 |

**Supplementary Table 5:** Results of sensitivity analysis in participants with any diabetes (N=4667).

|  | **Base Model** | | **Adjusted Model 1** | | **Fully Adjusted Model 2** | |
| --- | --- | --- | --- | --- | --- | --- |
| **Sleep Onset Time** | **Hazard Ratio (95% CI)** | ***p*-value** | **Hazard Ratio (95% CI)** | ***p*-value** | **Hazard Ratio (95% CI)** | ***p*-value** |
| **≥12:00am** | 1·29 (1·05-1·59) | 0·02 | 1·25 (1·01-1·55) | 0·04 | 1·23 (1·00-1·52) | 0·05 |
| **11:00pm-11:59pm** | 1·10 (0·99-1·24) | 0·08 | 1·09 (0·97-1·22) | 0·14 | 1·11 (0·99-1·24) | 0·08 |
| **10:00pm-10:59pm** | 1 (ref) | .. | 1 (ref) | .. | 1 (ref) | .. |
| **<10:00pm** | 1·31 (1·17-1·47) | < 0·005 | 1·22 (1·08-1·37) | < 0·005 | 1·22 (1·08-1·38) | < 0·005 |
| **Sleep Duration** | .. | .. | 0·94 (0·91-0·98) | < 0·005 | 0·97 (0·93-1·00) | 0·06 |
| **Sleep Irregularity** | .. | .. | 1·03 (1·00-1·06) | 0·03 | 1·01 (0·99-1·04) | 0·37 |

**Supplementary Table 6:** Results of sensitivity analysis in participants without any diabetes (N=83359).

|  | **Base Model** | | **Adjusted Model 1** | | **Fully Adjusted Model 2** | |
| --- | --- | --- | --- | --- | --- | --- |
| **Sleep Onset Time** | **Hazard Ratio (95% CI)** | ***p*-value** | **Hazard Ratio (95% CI)** | ***p*-value** | **Hazard Ratio (95% CI)** | ***p*-value** |
| **≥12:00am** | 0·63 (0·26-1·52) | 0·31 | 0·67 (0·28-1·63) | 0·38 | 0·80 (0·33-1·95) | 0·62 |
| **11:00pm-11:59pm** | 0·90 (0·57-1·40) | 0·63 | 0·84 (0·54-1·32) | 0·45 | 1·00 (0·62-1·59) | 0·99 |
| **10:00pm-10:59pm** | 1 (ref) | .. | 1 (ref) | .. | 1 (ref) | .. |
| **<10:00pm** | 1·37 (0·89-2·10) | 0·15 | 1·21 (0·77-1·89) | 0·41 | 1·39 (0·85-2·27) | 0·19 |
| **Sleep Duration** | .. | .. | 0·84 (0·74-0·95) | 0·01 | 0·95 (0·84-1·08) | 0·45 |
| **Sleep Irregularity** | .. | .. | 0·90 (0·79-1·02) | 0·09 | 0·95 (0·84-1·08) | 0·41 |

**Supplementary Table 7:** Results of sensitivity analysis in participants with hypotension (N=1018).

|  | **Base Model** | | **Adjusted Model 1** | | **Fully Adjusted Model 2** | |
| --- | --- | --- | --- | --- | --- | --- |
| **Sleep Onset Time** | **Hazard Ratio (95% CI)** | ***p*-value** | **Hazard Ratio (95% CI)** | ***p*-value** | **Hazard Ratio (95% CI)** | ***p*-value** |
| **≥12:00am** | 1·35 (1·07-1·71) | 0·01 | 1·27 (1·00-1·61) | 0·05 | 1·22 (0·96-1·54) | 0·11 |
| **11:00pm-11:59pm** | 1·15 (1·01-1·31) | 0·03 | 1·14 (1·01-1·30) | 0·04 | 1·15 (1·01-1·30) | 0·04 |
| **10:00pm-10:59pm** | 1 (ref) | .. | 1 (ref) | .. | 1 (ref) | .. |
| **<10:00pm** | 1·35 (1·19-1·54) | < 0·005 | 1·26 (1·10-1·44) | < 0·005 | 1·23 (1·07-1·41) | < 0·005 |
| **Sleep Duration** | .. | .. | 0·97 (0·93-1·01) | 0·13 | 1·00 (0·96-1·04) | 0·89 |
| **Sleep Irregularity** | .. | .. | 1·06 (1·03-1·09) | < 0·005 | 1·04 (1·01-1·07) | 0·01 |

**Supplementary Table 8:** Results of sensitivity analysis in participants without hypotension (N=75426).

|  | **Base Model** | | **Adjusted Model 1** | | **Fully Adjusted Model 2** | |
| --- | --- | --- | --- | --- | --- | --- |
| **Sleep Onset Time** | **Hazard Ratio (95% CI)** | ***p*-value** | **Hazard Ratio (95% CI)** | ***p*-value** | **Hazard Ratio (95% CI)** | ***p*-value** |
| **≥12:00am** | 1·35 (1·10-1·65) | < 0·005 | 1·28 (1·04-1·57) | 0·02 | 1·22 (1·00-1·50) | 0·05 |
| **11:00pm-11:59pm** | 1·11 (0·99-1·24) | 0·07 | 1·09 (0·98-1·22) | 0·12 | 1·12 (1·00-1·26) | 0·04 |
| **10:00pm-10:59pm** | 1 (ref) | .. | 1 (ref) | .. | 1 (ref) | .. |
| **<10:00pm** | 1·37 (1·23-1·53) | < 0·005 | 1·24 (1·11-1·39) | < 0·005 | 1·24 (1·10-1·40) | < 0·005 |
| **Sleep Duration** | .. | .. | 0·94 (0·90-0·97) | < 0·005 | 0·97 (0·94-1·00) | 0·07 |
| **Sleep Irregularity** | .. | .. | 1·05 (1·02-1·07) | < 0·005 | 1·02 (1·00-1·05) | 0·07 |

**Supplementary Table 9:** Results of sensitivity analysis in participants with HDL >1 (N=82645).

|  | **Base Model** | | **Adjusted Model 1** | | **Fully Adjusted Model 2** | |
| --- | --- | --- | --- | --- | --- | --- |
| **Sleep Onset Time** | **Hazard Ratio (95% CI)** | ***p*-value** | **Hazard Ratio (95% CI)** | ***p*-value** | **Hazard Ratio (95% CI)** | ***p*-value** |
| **≥12:00am** | 1·45 (0·76-2·75) | 0·26 | 1·43 (0·75-2·75) | 0·28 | 1·39 (0·72-2·68) | 0·32 |
| **11:00pm-11:59pm** | 1·13 (0·79-1·61) | 0·52 | 1·11 (0·78-1·59) | 0·56 | 1·13 (0·79-1·62) | 0·5 |
| **10:00pm-10:59pm** | 1 (ref) | .. | 1 (ref) | .. | 1 (ref) | .. |
| **<10:00pm** | 1·35 (0·96-1·90) | 0·09 | 1·25 (0·87-1·78) | 0·22 | 1·24 (0·85-1·79) | 0·26 |
| **Sleep Duration** | .. | .. | 0·92 (0·84-1·01) | 0·09 | 0·96 (0·87-1·06) | 0·38 |
| **Sleep Irregularity** | .. | .. | 1·00 (0·92-1·08) | 0·99 | 0·98 (0·90-1·06) | 0·57 |

**Supplementary Table 10:** Results of sensitivity analysis in participants with an HDL ≤1 (N=5381).

|  | **Base Model** | | **Adjusted Model 1** | | **Fully Adjusted Model 2** | |
| --- | --- | --- | --- | --- | --- | --- |
| **Sleep Onset Time** | **Hazard Ratio (95% CI)** | ***p*-value** | **Hazard Ratio (95% CI)** | ***p*-value** | **Hazard Ratio (95% CI)** | ***p*-value** |
| **≥12:00am** | 1·67 (1·24-2·25) | < 0·005 | 1·69 (1·25-2·29) | < 0·005 | 1·63 (1·20-2·21) | < 0·005 |
| **11:00pm-11:59pm** | 1·21 (1·02-1·44) | 0·03 | 1·17 (0·98-1·39) | 0·08 | 1·18 (0·99-1·40) | 0·07 |
| **10:00pm-10:59pm** | 1 (ref) | .. | 1 (ref) | .. | 1 (ref) | .. |
| **<10:00pm** | 1·57 (1·32-1·86) | < 0·005 | 1·40 (1·17-1·68) | < 0·005 | 1·34 (1·11-1·61) | < 0·005 |
| **Sleep Duration** | .. | .. | 0·88 (0·84-0·93) | < 0·005 | 0·92 (0·87-0·97) | < 0·005 |
| **Sleep Irregularity** | .. | .. | 0·99 (0·95-1·03) | 0·61 | 0·98 (0·94-1·02) | 0·26 |

**Supplementary Table 11:** Results of sensitivity analysis in female participants only (N=51214).

|  | **Base Model** | | **Adjusted Model 1** | | **Fully Adjusted Model 2** | |
| --- | --- | --- | --- | --- | --- | --- |
| **Sleep Onset Time** | **Hazard Ratio (95% CI)** | ***p*-value** | **Hazard Ratio (95% CI)** | ***p*-value** | **Hazard Ratio (95% CI)** | ***p*-value** |
| **≥12:00am** | 1·18 (0·92-1·52) | 0·2 | 1·08 (0·84-1·39) | 0·56 | 1·05 (0·81-1·35) | 0·73 |
| **11:00pm-11:59pm** | 1·05 (0·92-1·20) | 0·47 | 1·05 (0·91-1·20) | 0·52 | 1·08 (0·94-1·24) | 0·27 |
| **10:00pm-10:59pm** | 1 (ref) | .. | 1 (ref) | .. | 1 (ref) | .. |
| **<10:00pm** | 1·26 (1·11-1·44) | < 0·005 | 1·15 (1·00-1·32) | 0·05 | 1·17 (1·01-1·35) | 0·03 |
| **Sleep Duration** | .. | .. | 0·96 (0·92-1·00) | 0·04 | 0·99 (0·95-1·03) | 0·77 |
| **Sleep Irregularity** | .. | .. | 1·08 (1·04-1·11) | < 0·005 | 1·05 (1·02-1·08) | < 0·005 |

**Supplementary Table 12:** Results of sensitivity analysis in male participants only (N=36812).

|  | **Base Model** | | **Adjusted Model 1** | | **Fully Adjusted Model 2** | |
| --- | --- | --- | --- | --- | --- | --- |
| **Sleep Onset Time** | **Hazard Ratio (95% CI)** | ***p*-value** | **Hazard Ratio (95% CI)** | ***p*-value** | **Hazard Ratio (95% CI)** | ***p*-value** |
| **≥12:00am** | 1·95 (0·62-6·12) | 0·25 | 1·77 (0·56-5·64) | 0·33 | 1·94 (0·61-6·18) | 0·26 |
| **11:00pm-11:59pm** | 1·45 (0·73-2·89) | 0·29 | 1·39 (0·69-2·77) | 0·35 | 1·51 (0·75-3·03) | 0·24 |
| **10:00pm-10:59pm** | 1 (ref) | .. | 1 (ref) | .. | 1 (ref) | .. |
| **<10:00pm** | 2·43 (1·24-4·74) | 0·01 | 1·93 (0·96-3·87) | 0·07 | 1·99 (0·96-4·12) | 0·06 |
| **Sleep Duration** | .. | .. | 0·83 (0·70-0·98) | 0·02 | 0·86 (0·72-1·02) | 0·08 |
| **Sleep Irregularity** | .. | .. | 1·04 (0·92-1·18) | 0·5 | 1·04 (0·91-1·19) | 0·56 |

**Supplementary Table 13:** Results of sensitivity analysis in participants aged 50 or under (N=9106).

|  | **Base Model** | | **Adjusted Model 1** | | **Fully Adjusted Model 2** | |
| --- | --- | --- | --- | --- | --- | --- |
| **Sleep Onset Time** | **Hazard Ratio (95% CI)** | ***p*-value** | **Hazard Ratio (95% CI)** | ***p*-value** | **Hazard Ratio (95% CI)** | ***p*-value** |
| **≥12:00am** | 1·35 (0·96-1·90) | 0·09 | 1·30 (0·92-1·84) | 0·14 | 1·23 (0·87-1·74) | 0·24 |
| **11:00pm-11:59pm** | 1·24 (1·02-1·50) | 0·03 | 1·22 (1·01-1·48) | 0·04 | 1·26 (1·04-1·53) | 0·02 |
| **10:00pm-10:59pm** | 1 (ref) | .. | 1 (ref) | .. | 1 (ref) | .. |
| **<10:00pm** | 1·48 (1·22-1·79) | < 0·005 | 1·36 (1·12-1·66) | < 0·005 | 1·35 (1·10-1·66) | < 0·005 |
| **Sleep Duration** | .. | .. | 0·94 (0·88-0·99) | 0·03 | 0·97 (0·92-1·03) | 0·36 |
| **Sleep Irregularity** | .. | .. | 1·03 (0·99-1·08) | 0·17 | 1·01 (0·96-1·05) | 0·76 |

**Supplementary Table 14:** Results of sensitivity analysis in participants aged between 50 and 64 (N=40746).

|  | **Base Model** | | **Adjusted Model 1** | | **Fully Adjusted Model 2** | |
| --- | --- | --- | --- | --- | --- | --- |
| **Sleep Onset Time** | **Hazard Ratio (95% CI)** | ***p*-value** | **Hazard Ratio (95% CI)** | ***p*-value** | **Hazard Ratio (95% CI)** | ***p*-value** |
| **≥12:00am** | 1·33 (1·05-1·69) | 0·02 | 1·27 (1·00-1·61) | 0·05 | 1·23 (0·96-1·56) | 0·09 |
| **11:00pm-11:59pm** | 1·04 (0·92-1·19) | 0·51 | 1·03 (0·90-1·17) | 0·68 | 1·05 (0·92-1·20) | 0·46 |
| **10:00pm-10:59pm** | 1 (ref) | .. | 1 (ref) | .. | 1 (ref) | .. |
| **<10:00pm** | 1·30 (1·14-1·48) | < 0·005 | 1·17 (1·03-1·34) | 0·02 | 1·17 (1·02-1·35) | 0·02 |
| **Sleep Duration** | .. | .. | 0·94 (0·90-0·97) | < 0·005 | 0·97 (0·93-1·01) | 0·13 |
| **Sleep Irregularity** | .. | .. | 1·05 (1·02-1·08) | < 0·005 | 1·02 (0·99-1·06) | 0·12 |

**Supplementary Table 15:** Results of sensitivity analysis in participants aged greater than 64 (N=38174).

|  | **Base Model** | | **Adjusted Model 1** | | **Fully Adjusted Model 2** | |
| --- | --- | --- | --- | --- | --- | --- |
| **Sleep Onset Time** | **Hazard Ratio (95% CI)** | ***p*-value** | **Hazard Ratio (95% CI)** | ***p*-value** | **Hazard Ratio (95% CI)** | ***p*-value** |
| **≥12:00am** | 1·36 (1·10-1·68) | < 0·005 | 1·29 (1·04-1·60) | 0·02 | 1·25 (1·01-1·55) | 0·04 |
| **11:00pm-11:59pm** | 1·11 (0·99-1·25) | 0·08 | 1·09 (0·97-1·23) | 0·15 | 1·11 (0·99-1·25) | 0·08 |
| **10:00pm-10:59pm** | 1 (ref) | .. | 1 (ref) | .. | 1 (ref) | .. |
| **<10:00pm** | 1·35 (1·20-1·52) | < 0·005 | 1·20 (1·07-1·36) | < 0·005 | 1·19 (1·05-1·35) | 0·01 |
| **Sleep Duration** | .. | .. | 0·92 (0·89-0·95) | < 0·005 | 0·95 (0·92-0·99) | 0·01 |
| **Sleep Irregularity** | .. | .. | 1·05 (1·02-1·07) | < 0·005 | 1·02 (0·99-1·05) | 0·12 |

**Supplementary Table 16:** Results of sensitivity analysis for time to CVD event, discounting the first 12 months after accelerometer measurement (N=87312).

|  | **Base Model** | | **Adjusted Model 1** | | **Fully Adjusted Model 2** | |
| --- | --- | --- | --- | --- | --- | --- |
| **Sleep Onset Time** | **Hazard Ratio (95% CI)** | ***p*-value** | **Hazard Ratio (95% CI)** | ***p*-value** | **Hazard Ratio (95% CI)** | ***p*-value** |
| **≥12:00am** | 1·26 (1·00-1·59) | 0·05 | 1·20 (0·95-1·52) | 0·12 | 1·16 (0·92-1·47) | 0·21 |
| **11:00pm-11:59pm** | 1·12 (0·99-1·27) | 0·07 | 1·10 (0·97-1·25) | 0·14 | 1·12 (0·99-1·27) | 0·08 |
| **10:00pm-10:59pm** | 1 (ref) | .. | 1 (ref) | .. | 1 (ref) | .. |
| **<10:00pm** | 1·34 (1·18-1·52) | < 0·005 | 1·19 (1·04-1·35) | 0·01 | 1·16 (1·01-1·33) | 0·03 |
| **Sleep Duration** | .. | .. | 0·91 (0·88-0·95) | < 0·005 | 0·95 (0·91-0·98) | 0·01 |
| **Sleep Irregularity** | .. | .. | 1·04 (1·01-1·07) | < 0·005 | 1·02 (0·99-1·05) | 0·2 |

**Supplementary Table 17:** Results of sensitivity analyses for time to CVD event, discounting the first 18 months after accelerometer measurement (N=86905).
